# Leveraging functional annotations to map rare variants associated with Alzheimer’s disease with gruyere

**DOI:** 10.1101/2024.12.06.24318577

**Authors:** Anjali Das, Chirag Lakhani, Chloé Terwagne, Jui-Shan T. Lin, Tatsuhiko Naito, Towfique Raj, David A. Knowles

## Abstract

The increasing availability of whole-genome sequencing (WGS) has begun to elucidate the contribution of rare variants (RVs), both coding and non-coding, to complex disease. Multiple RV association tests are available to study the relationship between genotype and phenotype, but most are restricted to per-gene models and do not fully leverage the availability of variant-level functional annotations. We propose Genome-wide Rare Variant EnRichment Evaluation (`gruyere`), a Bayesian probabilistic model that complements existing methods by learning global, trait-specific weights for functional annotations to improve variant prioritization. We apply `gruyere` to WGS data from the Alzheimer’s Disease (AD) Sequencing Project, consisting of 7,966 cases and 13,412 controls, to identify AD-associated genes and annotations. Growing evidence suggests that disruption of microglial regulation is a key contributor to AD risk, yet existing methods have not had sufficient power to examine rare non-coding effects that incorporate such cell-type specific information. To address this gap, we 1) use predicted enhancer and promoter regions in microglia and other potentially relevant cell types (oligodendrocytes, astrocytes, and neurons) to define per-gene non-coding RV test sets and 2) include cell-type specific variant effect predictions (VEPs) as functional annotations. `gruyere` identifies 15 significant genetic associations not detected by other RV methods and finds deep learning-based VEPs for splicing, transcription factor binding, and chromatin state are highly predictive of functional non-coding RVs. Our study establishes a novel and robust framework incorporating functional annotations, coding RVs, and cell-type associated non-coding RVs, to perform genome-wide association tests, uncovering AD-relevant genes and annotations.

## 1 Main

The recent increase in available whole-genome sequencing (WGS) data has facilitated the study of rare variants (RVs), particularly in understanding their effects on complex diseases like late-onset Alzheimer’s disease (AD). AD is a neurodegenerative disorder with an estimated heritability between 59% and 74% [1]. While genome-wide association studies (GWAS) have identified over 100 loci linked to AD, with the *APOE-e4* allele as the strongest genetic risk factor, they are restricted to common variant associations [2, 3]. Despite considerable efforts to quantify the polygenic nature of AD, a significant portion of genetic heritability remains unaccounted for. Some of this missing heritability may be recovered with RVs [4]. RVs generally exhibit larger effect sizes than common variants, but their role is not yet well understood [5]. Studies have shown that integrating RVs into cumulative polygenic risk scores (PRS) can enhance predictive performance [6], but existing methods have identified fewer gene associations and have lower predictive power compared to common variant approaches. While a number of genes, including *TREM2, ABCA7* and *SORL1* [7], have known RV associations in AD, the majority of these findings are restricted to coding variants. As most GWAS signals lie in the non-coding genome, expanding RV association studies beyond coding variants is critical. However, the study of non-coding RVs poses challenges due to the vast number of these variants, most of which likely have no functional impact [8]. It is therefore of substantial interest to use functional annotations for variant filtering and prioritization. To develop a more robust understanding of the contributions of both coding and non-coding RVs to AD, we propose a novel method that not only weights variants according to annotations but also prioritizes functional annotations that are most trait-relevant.

Applying traditional variant-level approaches like GWAS to RVs has low statistical power due to sparsity and a high multiple testing burden due to the large number of RVs compared to common variants. To address these limitations, RV methods aggregate variants in biologically related regions, typically by gene, to increase power [9]. More recent RV methods additionally account for functional annotations to prioritize relevant variants and filter out those predicted to have no function, which otherwise reduce power [10, 11]. Despite growing efforts to accurately predict which variants will affect particular molecular phenotypes (e.g., enhancer activation, RNA splicing) [12– 16], there is a limited understanding of *which functions* are the most disease-relevant. Using functional annotations that have no phenotypic associations to weight RVs can add noise to models and decrease their power. This motivated us to develop a method that learns a genome-wide mapping from functional annotations to variant importance.

Growing evidence suggests that disrupted gene regulation in central nervous system (CNS) cell types, particularly microglia, is associated with the development and progression of AD [1, 17]. The majority of RV tests are developed for coding variant associations because 1) predicting functional coding variants is comparatively straight-forward (at least for loss-of-function), 2) population-scale whole-exome sequencing predates WGS, and 3) defining non-coding regions for testing is challenging in itself. Some methods use sliding windows, but testing overlapping windows of varying sizes can result in loss of power due to multiple testing [18]. Other methods use predicted *cis*-regulatory elements (CREs), in particular enhancers and promoters, to construct testing regions [19–21]. Given their modest size (typically less than 2kb), testing individual CREs still has limited statistical power. Combining multiple CREs that regulate a gene could help address this limitation but relies on accurate predictions of enhancer-gene links. We leverage the Activity-by-Contact (ABC) model, which predicts cell-type specific enhancer-gene connectivity using chromatin state and conformation data [22]. We aggregate ABC-predicted enhancer-gene pairs to determine non-coding, cell-type and gene specific RV testing regions.

Due to the large number of genes and several million RVs found in population-scale WGS, existing methods are primarily restricted to per-gene models. This limits our understanding of disease-associated functional annotations. Most existing RV methods are explicitly, or can be viewed as, generalized linear mixed models (GLMM). We instead develop a Bayesian generalized linear model (GLM) Genome-wide Rare Variant EnRichment Evaluation, or `gruyere`, to model cell-type specific, non-coding RV associations on a genome-wide scale. In `gruyere`, a variant’s effect is a deterministic function of its annotations and the estimated AD-relevance of the gene it is linked to (if any). Our Bayesian model iteratively learns AD-relevant gene effects, covariate weights, and functional annotation importance while quantifying uncertainty, providing increased flexibility to capture the complex, hierarchical structure of genetic data. We test our model using simulation analyses and compare results to several existing RV methods. We apply `gruyere` to WGS data from the Alzheimer’s Disease Sequencing Project (ADSP), consisting of 7,966 cases and 13,412 controls. Our model determines splicing, transcription factor (TF) binding, and chromatin state annotations most enriched for AD-associated non-coding RVs and identifies 16 significant genes, 15 of which are uniquely identified by `gruyere`. Of these, four – *C9orf78, MAF1, NUP93*, and *GALNT9* – remain significant in omnibus tests.

## 2 Results

### 2.1 Genome-wide Rare Variant EnRichment Evaluation (gruyere) overview

**Table 1.**
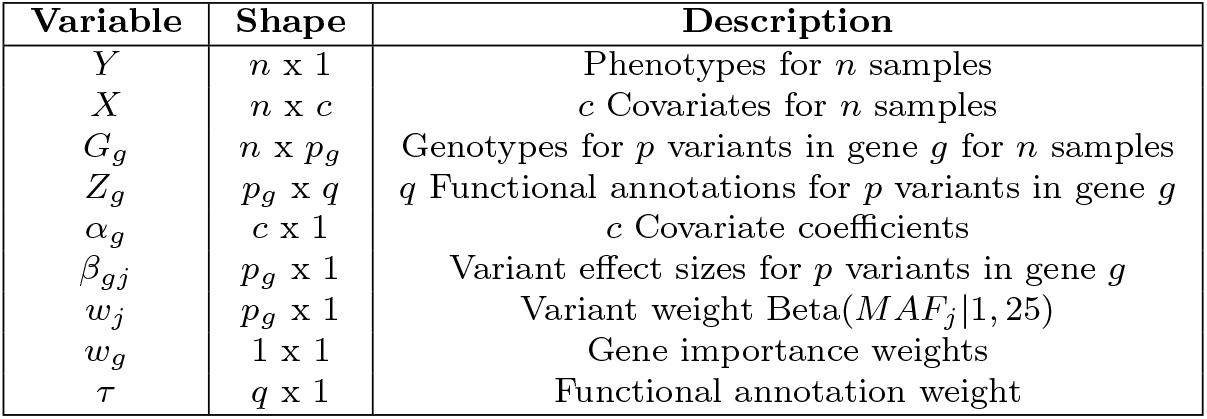
Summary of gruyere Variables.

Current RV methods rely on independent per-gene models and, therefore, cannot capture genome-wide functional annotation importance. `gruyere` serves as a complementary method to existing RV tests by learning trait-specific functional annotation weights, covariate coefficients, and gene effects under a Bayesian framework (Supplemental Figure 1, Table 1). Rather than modeling each gene separately, we jointly fit `gruyere` as a hierarchy of per-gene GLMs using stochastic variational inference (SVI) [23]. We model AD risk for each gene *g*,

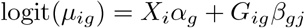

where *µ*_*ig*_ is the probability of AD for individual *i* given the genotypes for RVs associated with gene *g, X*_*i*_ is a vector of covariates (e.g. sex, age, *APOE-e4* genotype), and *G*_*ig*_ is a genotype vector. We learn covariate weights *α*_*g*_ and variant effects *β*_*gj*_. We set *β*_*gj*_ to be a deterministic function of a learned gene effect *w*_*g*_, transformed minor allele frequencies (MAFs) *w*_*j*_, functional annotations *Z* (detailed in Methods 3.4 and Figure 1C), and learned annotation importance weights *τ*,

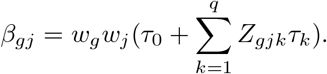

**Fig. 1.**
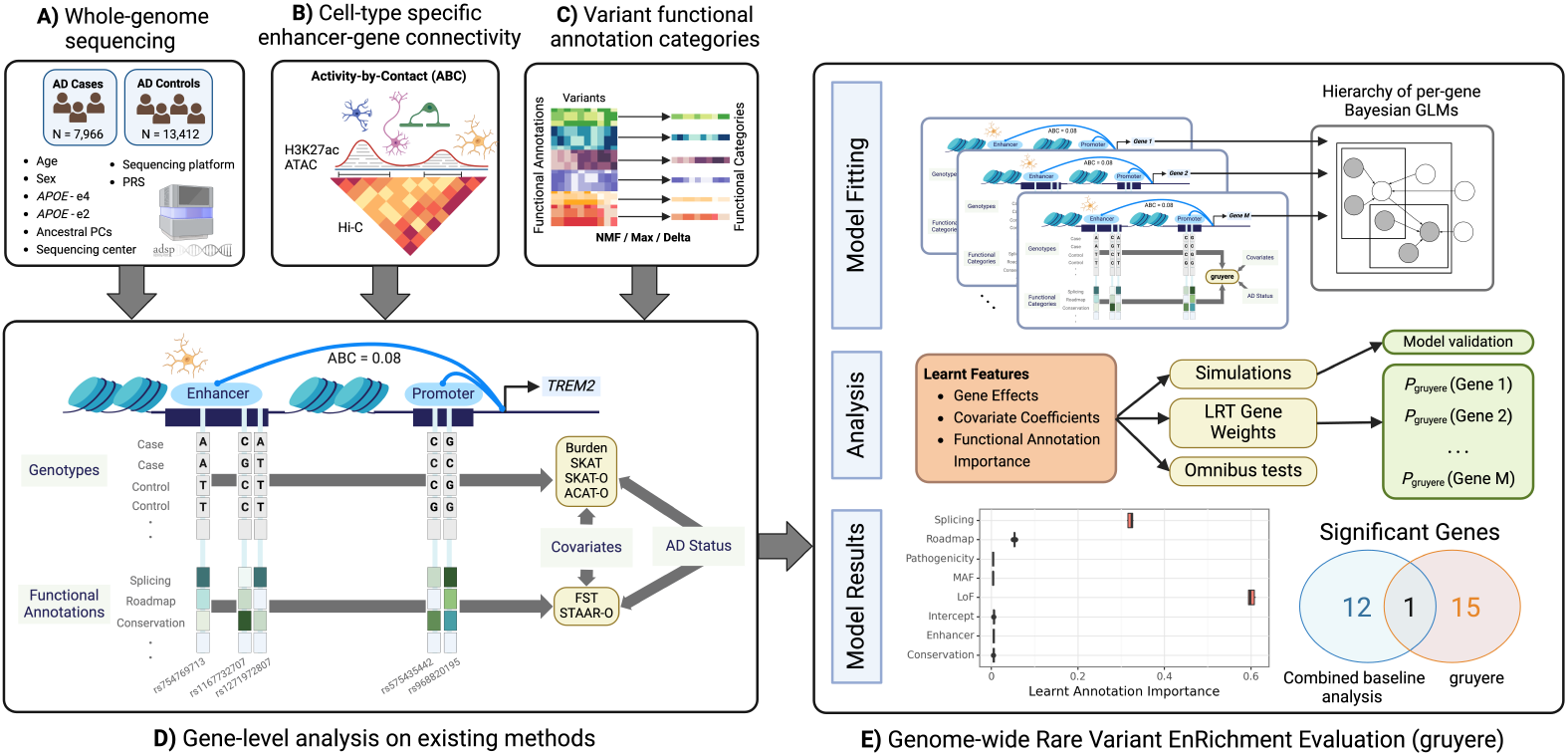
Overview of the application of gruyere to AD. Input data includes **A)** WGS and clinical information for AD cases and controls, **B)** Enhancer-gene interactions predicted by the ABC model for microglia, oligodendrocytes, astrocytes, and neurons, and **C)** variant functional annotations. **D)** Example analysis for the *TREM2* gene and microglia cell-type on existing methods. Columns represent RVs; light grey rectangles represent individual-level genotypes from WGS data for cases and controls; functional annotations for each RV are shown below genotypes; Burden, SKAT, SKAT-O, and ACAT-O are existing tests that use genotype, covariate, and AD status; FST and STAAR-O additionally use functional annotations. **E)** Workflow for `gruyere`. Per-gene RVs are aggregated and used for fitting the hierarchical Bayesian GLM. `gruyere` learns weights for covariates, genes, and functional annotations. We use simulations to assess `gruyere` at different heritabilities. Likelihood ratio tests are used to calculate gene-level *p*-values. Optionally, the `gruyere` *p*-values can be integrated with existing methods through omnibus testing.

In our analyses, `gruyere` learns annotation weights *τ* for a range of annotations *Z*, including in silico mutagenesis deep learning model predictions of splicing disruption (derived as the maximum of four individual SpliceAI scores [24]) and cell-type specific TF binding and chromatin state (derived from the Enformer model [14]). A larger magnitude of *w*_*g*_ indicates that disruption of gene *g* is associated with a higher predicted risk of AD. Similar to a burden test, `gruyere` assumes all variants within a gene have the same direction of effect [25, 26]. However, because our functional annotations include both loss- and gain-of-function predictions, we are able to capture additional dispersion-based signal. To ensure robust generalization of learned parameters, we split data into training (80%) and test (20%) sets, where model weights are optimized using the training set and assessed on the unseen test set. We apply `gruyere` to both coding and non-coding RVs for AD, defining four cell-type specific non-coding groups for AD-relevant cell types (microglia, oligodendrocytes, astrocytes, and neurons [17]) and testing each group individually.

#### Step 1. Estimating global annotation weights τ

Fitting *τ* jointly across the entire genome would be 1) computationally challenging due to the large number of RVs and 2) statistically inefficient, as only AD-associated genes will contribute relevant signal. We therefore estimate *τ* under the `gruyere` model from a subset of potentially AD-relevant genes identified using a lenient significance threshold (nominal *p <* 0.01) for the Functional Score Test (FST)[27]. We assess the robustness of `gruyere` estimates when selecting genes with varied significance thresholds and for a number of existing RV tests and find annotation weights *τ* are broadly consistent (+*/* − 0.02).

#### Step 2. Per-gene analysis

Once genome-wide estimates for *τ* are obtained, `gruyere` simplifies to a logistic regression that learns covariate *α*_*g*_ and gene *w*_*g*_ weights. We efficiently fit `gruyere` separately and in parallel for all genes, holding *τ* fixed. We perform likelihood ratio tests (LRT) to compare a covariate-only regression against combined covariate and genotype regression models to determine gene-level significance for *w*_*g*_.

### 2.2 Constructing cell-type & gene specific variant sets using predicted CREs

We grouped non-coding RVs by gene and CNS cell type using the Activity-by-Contact (ABC) model (Figure 1B) [22]. The ABC model uses epigenomic profiles and chromatin conformation to determine cell-type specific enhancer-gene interactions, filtering out genes that are not expressed. We use publicly available ATAC-seq and H3K27ac ChIP-seq signals for microglia, oligodendrocytes, astrocytes, and neurons [28], as well as Hi-C averaged across ten CNS cell types to account for 3-dimensional chromatin interactions. For each gene, we analyze RVs aggregated across all CREs interacting with that gene (*ABC >* 0.02). We test each cell type separately and also analyze rare coding variants for comparison. In total, ABC defines 70,300 CREs across all four cell types and 17,929 genes, with higher relative counts of microglia-predicted enhancers (Figure 2A). Predicted CREs frequently co-occur across cell types, with 39.4% of CREs found in more than one cell type. Promoter regions tend to have higher ABC scores than enhancers (mean *ABC* = 0.07 vs. mean *ABC* = 0.04), but their genomic lengths are similar, with an average length of 632bp and standard deviation of 132bp. ABC accounts for interactions of a single enhancer with multiple genes so one RV can be linked to multiple genes. In our analysis, an ABC enhancer maps to an average of 3.8 genes in microglia and between 5.4 and 5.9 genes in the other three cell types. Our non-coding variant sets contain an average of 376 RVs per gene. There are a total of 2,092,931 RVs in CREs across the four cell types, 901,570 of which are included for more than one cell type, and 550,001 that are in all four cell types (Figure 2B).

**Fig. 2.**
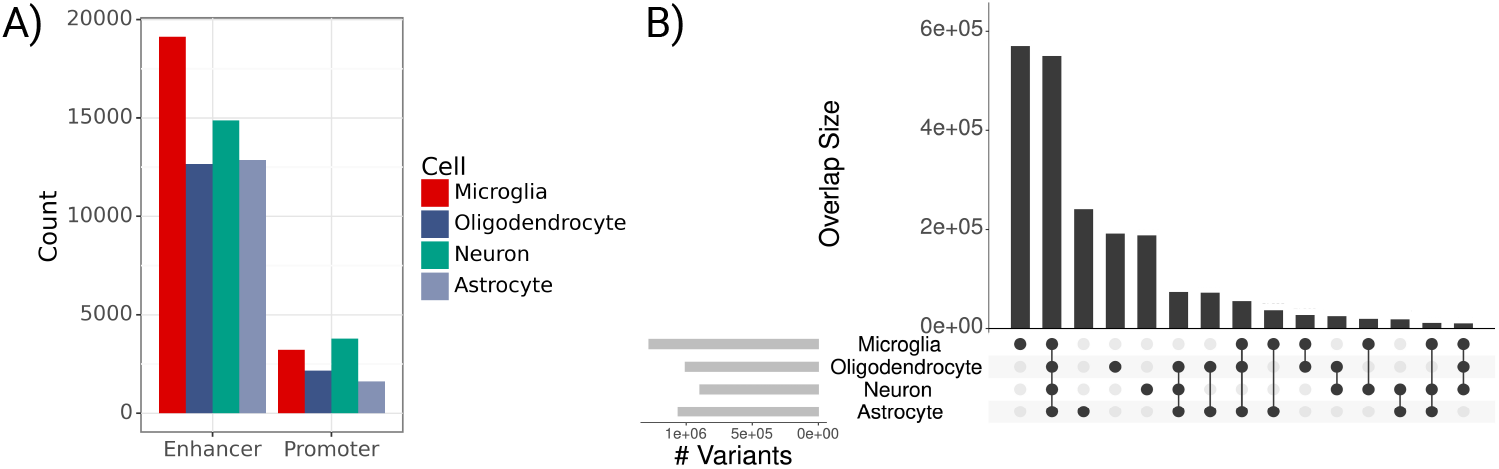
Predicted regulatory element and variant counts across cell types. **A)** Bar plot of predicted CRE counts by cell type (*ABC >* 0.02). **B)** Upset plot of variant overlap across 4 cell types in ADSP data; Light grey bars on left indicate total RV counts for each cell type; Vertically connected dots represents groups and corresponding bars indicate variant overlap for that group.

### 2.3 Simulation studies confirm accurate estimation of model parameters

We generate synthetic phenotypes (see Methods 3.5) and fit `gruyere` on 100 sets of simulated data with estimated heritability between 5% and 30% (detailed in Supplemental Methods [29]) using 500 randomly selected genes. We find that all variables are well recovered, with average Pearson correlations *R* = 0.81, 0.95, 0.98, 0.97 for *α*_*g*_, *β*_*gj*_, *w*_*g*_ and *τ* respectively (Figure 3). Covariates *α*_*g*_ have the lowest *R*, possibly due to correlated covariates. Average recovery across all variables remains high when varying the prior distributions (Pearson *R >* 0.78) as well as when simulated distributions differ from the priors used during inference (Pearson *R >* 0.66). Results are robust to the number of covariates, genes, and annotations modeled.

**Fig. 3.**
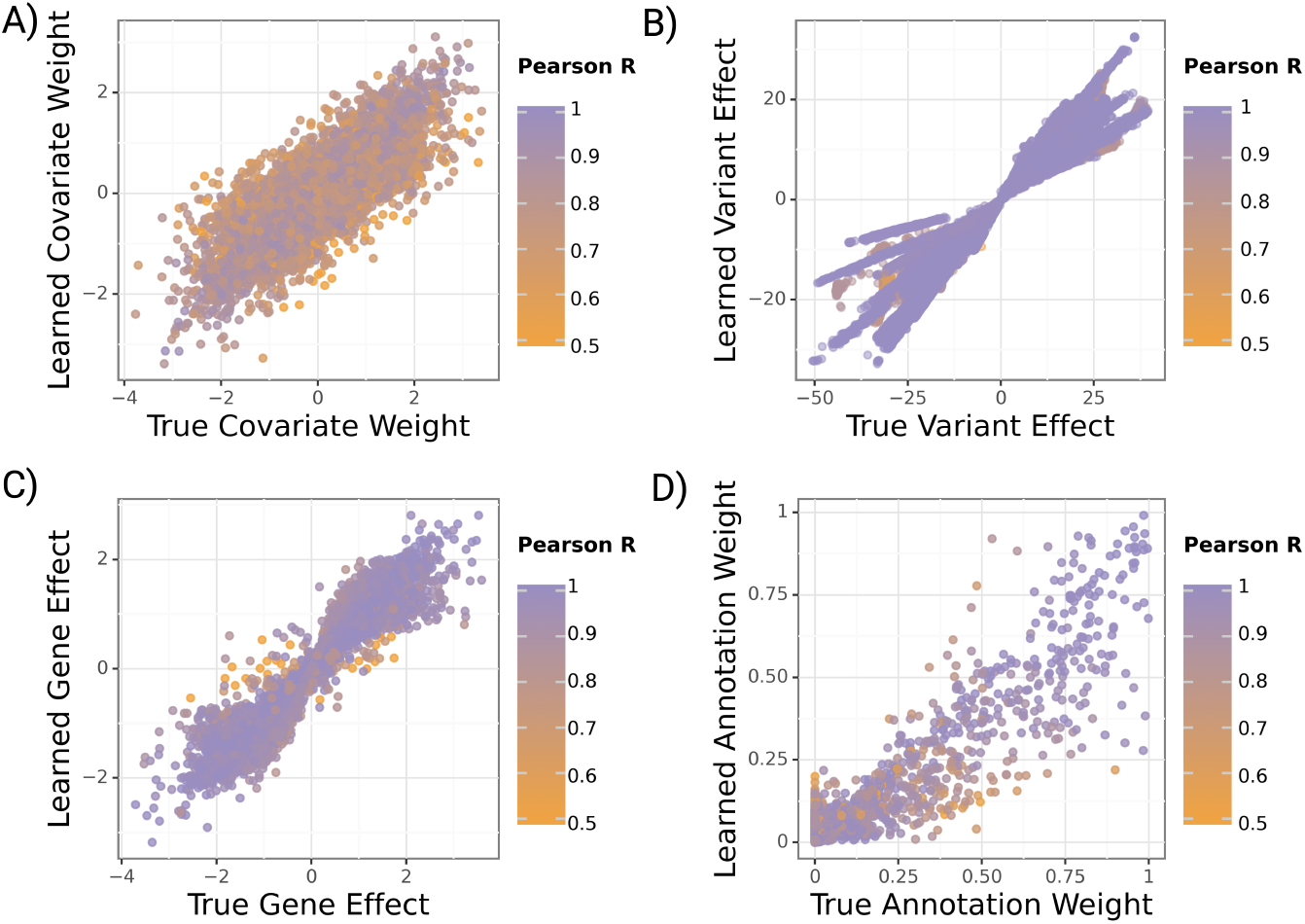
Learned versus true gruyere parameters across 100 simulations. Points are colored by the Pearson correlation coefficient of a parameter for a given simulation. **A)** Covariate regression coefficients *α* (*c* = 30 covariates). **B)** Variant effect *β*. **C)** Gene effect *w*_*g*_ (*M* = 500 randomly selected genes). **D)** Annotation weight *τ* (*q* = 13 annotations).

We analyze how simulation performance correlates with overall and genetic heritability for each simulation. This allows us to more meaningfully evaluate model performance for complex diseases like AD where estimated heritability is low. As expected, we find that `gruyere` is better able to recover *β*_*gj*_ and *w*_*g*_ with increased genetic heritability (Supplemental Figure 2). However, even when total heritability is as low as 5%, the minimum correlation between true and estimated parameters remains quite high (Pearson *R* = 0.68).

### 2.4 Applying gruyere to AD WGS data reveals novel disease associations

#### Performance on ADSP WGS data

After validating model performance through simulations, we fit `gruyere` to the ADSP WGS data. We analyze coding and non-coding (microglia, oligodendrocyte, neuron, astrocyte) groups separately, and refer to each set as a cell type. For each cell type, we use a subset of genes for joint fitting (FST *p*-value *<* 0.01), leading to between 267 and 333 genes per cell-type. AD prediction performance is fairly consistent across non-coding variants (average test set Area Under the Receiver Operating Characteristic, or AUROC, of 0.69) and slightly higher for coding variants (average *AUROC*_test_ = 0.70) (Figure 4A-B). When averaging predicted probabilities across genes, performance further improves (*AUROC*_test_ = 0.72) for all cell types. These metrics are in line with current AD literature and outperform a covariate-only regression model (*AUROC*_test_ = 0.65) [30]. There is a substantial range in prediction performance for each cell type (e.g. minimum *AUROC*_test_ = 0.62, maximum *AUROC*_test_ = 0.71 for microglia), highlighting the varying degrees of association with AD across genes. We find that gene-level performance is consistent across model refitting and that the loss converges reliably (4C) [31]. Fitting time increases approximately logarithmically with the total number of genes (Figure 4D). On average, it takes 37 seconds per epoch and three hours total to jointly fit `gruyere` across 300 epochs. Per-gene estimation is much faster, taking an average of 4.3 seconds per gene to complete.

**Fig. 4.**
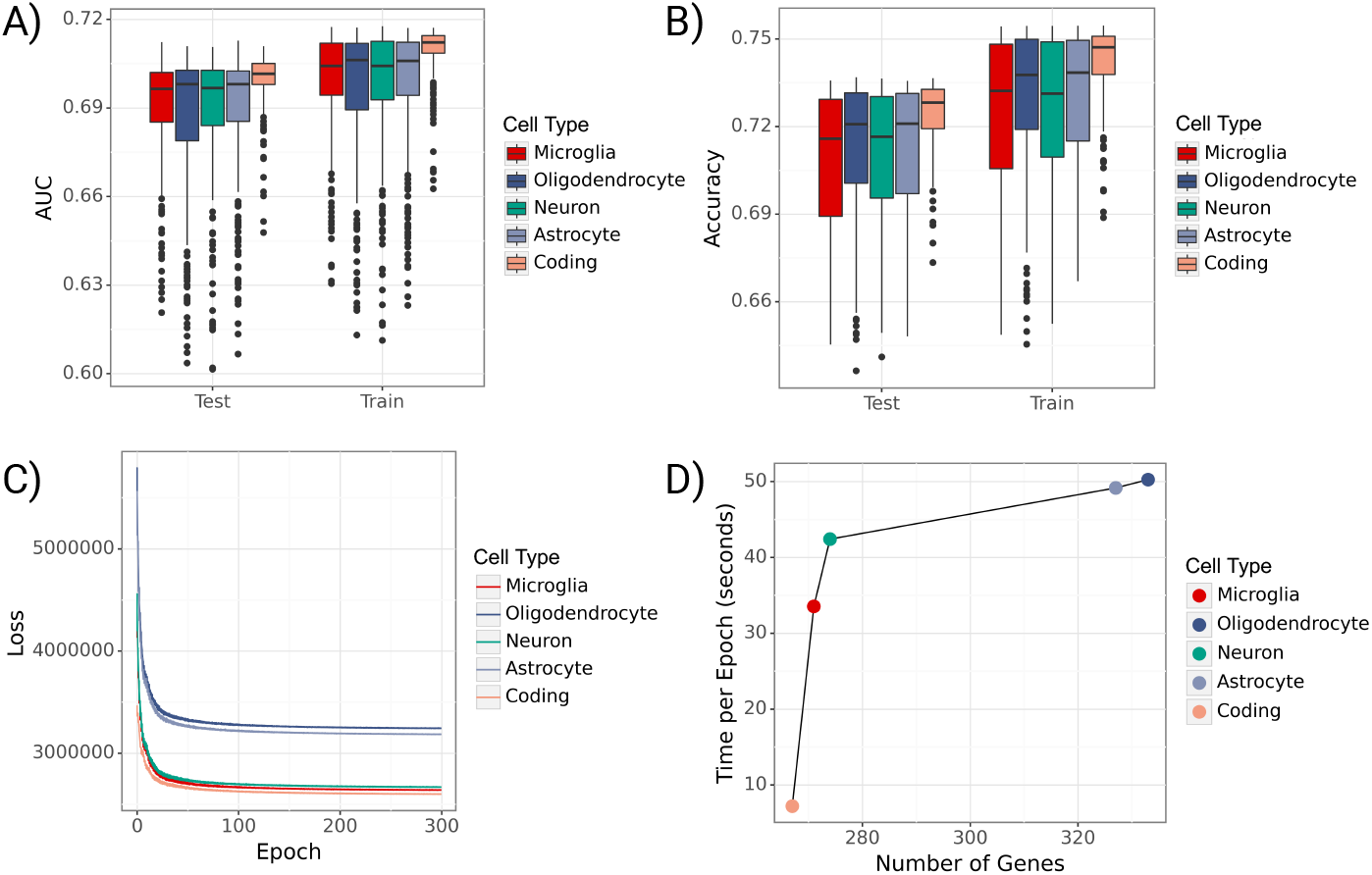
Performance of gruyere. **A)** Boxplots of per-gene AUROCs for train and test sets across cell types. **B)** Boxplots of per-gene accuracies for train and test sets across cell types. **C)** Trace ELBO loss over 300 epochs across cell types. **D)** Average training time per epoch (seconds) versus number of genes used in joint model for each cell type.

#### Learned annotation and covariate weights

We find that the top `gruyere` functional annotation weights come from splicing across all non-coding RV groups and loss-of-function (LoF) for coding variants (Figure 5A). LoF variants can be highly disruptive to gene function and are often used as a variant filtering method in gene-based tests. Therefore, it is predictable that we find `gruyere` places a large weight on LoF coding RVs. It is perhaps not surprising that `gruyere` also prioritizes RVs predicted to disrupt normal splicing, as they can substantially change the protein product or have large effects on gene dosage via nonsense mediated decay. For all non-coding regions, cell-type specific TF binding predictions from Enformer contain the next largest annotation weights. This suggests RVs associated with an increase (Max TF Delta) or decrease (Min TF Delta) in binding are predicted to have larger effects on AD, at least in AD-relevant genes. For microglia RV sets, we additionally find increased AD association for variants related to histone modification (H3K4me3, H3K27me3, H3K27ac) and DNASE in monocytes (often used as a proxy for microglia [32]). We restrict Enformer annotations to non-coding variants as they are specific to cell-types. The enhancer category has very small weights across cell-types. Since all variants included in the non-coding analyses are in putative CREs, it is perhaps not too surprising that cell-type specific enhancer annotations are lowly prioritized by `gruyere`.

**Fig. 5.**
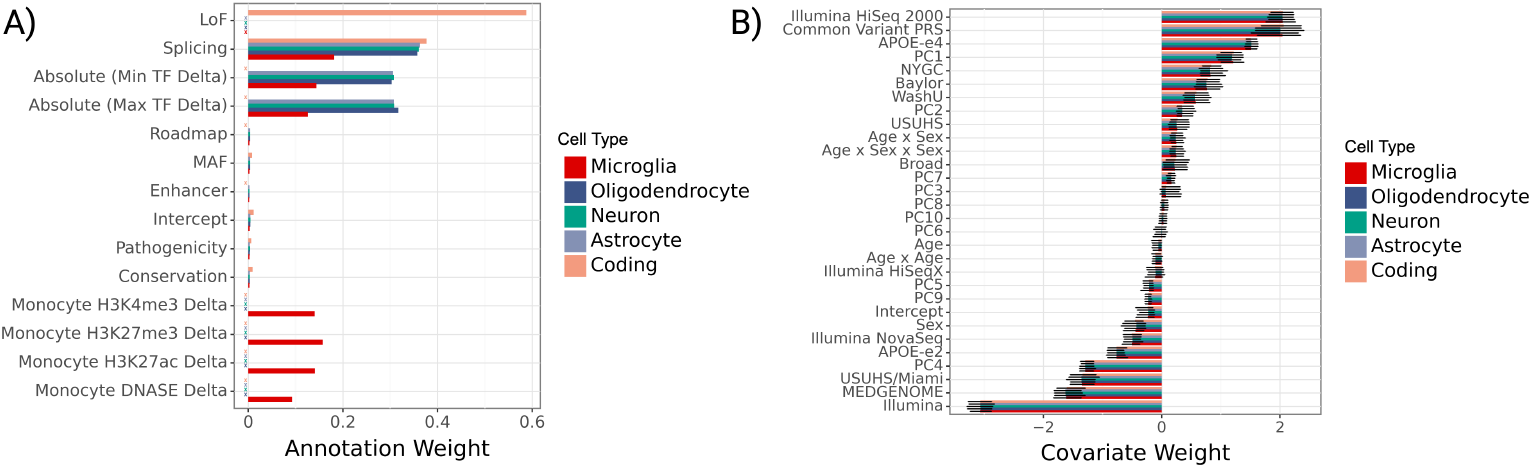
gruyere parameters learned from ADSP WGS. **A)** Bar plot of genome-wide annotation weights *τ* learned in jointly fit model across cell types. We denote crosses (X) to the left of bars where an annotation is not included for a cell type. **B)** Bar plot of per-gene covariate weights (*α*_*g*_) learned in jointly fit model across cell types. Error bars illustrate the minimum and maximum values learned across genes.

Covariate effects are learned consistently across genes and cell types, with sequencing center, common variant polygenic risk score, and Illumina HiSeq 2000 sequencing platform as the top three covariates (Figure 5B). As expected, *APOE-e4* is learned to have a large positive risk effect while the *APOE-e2* allele has a negative (protective) effect [2]. These effects agree well with those of a simple logistic regression predicting AD status from covariates.

#### Learned gene effects and associations

Estimated gene effects *w*_*g*_ are fit from a per-gene logistic regression, where we use LRTs to determine gene-level `gruyere` *p*-values (Methods 3.2). Significant genes after Bonferroni correction for each cell type are shown in Figure 6A, where a total of 16 genes reach genome-wide significance. The well-established *TREM2* RV association [33], as well as *MAF1, C9orf78*, and *GRIK3* are found significant for coding variants. Although not as widely recognized as *TREM2, MAF1* has been previously reported in association with AD [34], and *C9orf78* has been identified in an AD dementia meta-analysis [35]. *GRIK3* has emerged as a gene of interest due to the role of kainate receptors in neuroinflamation, a key feature of AD. Inflammatory responses can amplify glutamate release and disrupt receptor functioning, which may further accelerate neurodegeneration. This makes *GRIK3*, and glutamate signaling more broadly, potential targets for therapies [36, 37]. The identification of these genes by `gruyere` highlights their potential as candidate genes for further study in AD.

**Fig. 6.**
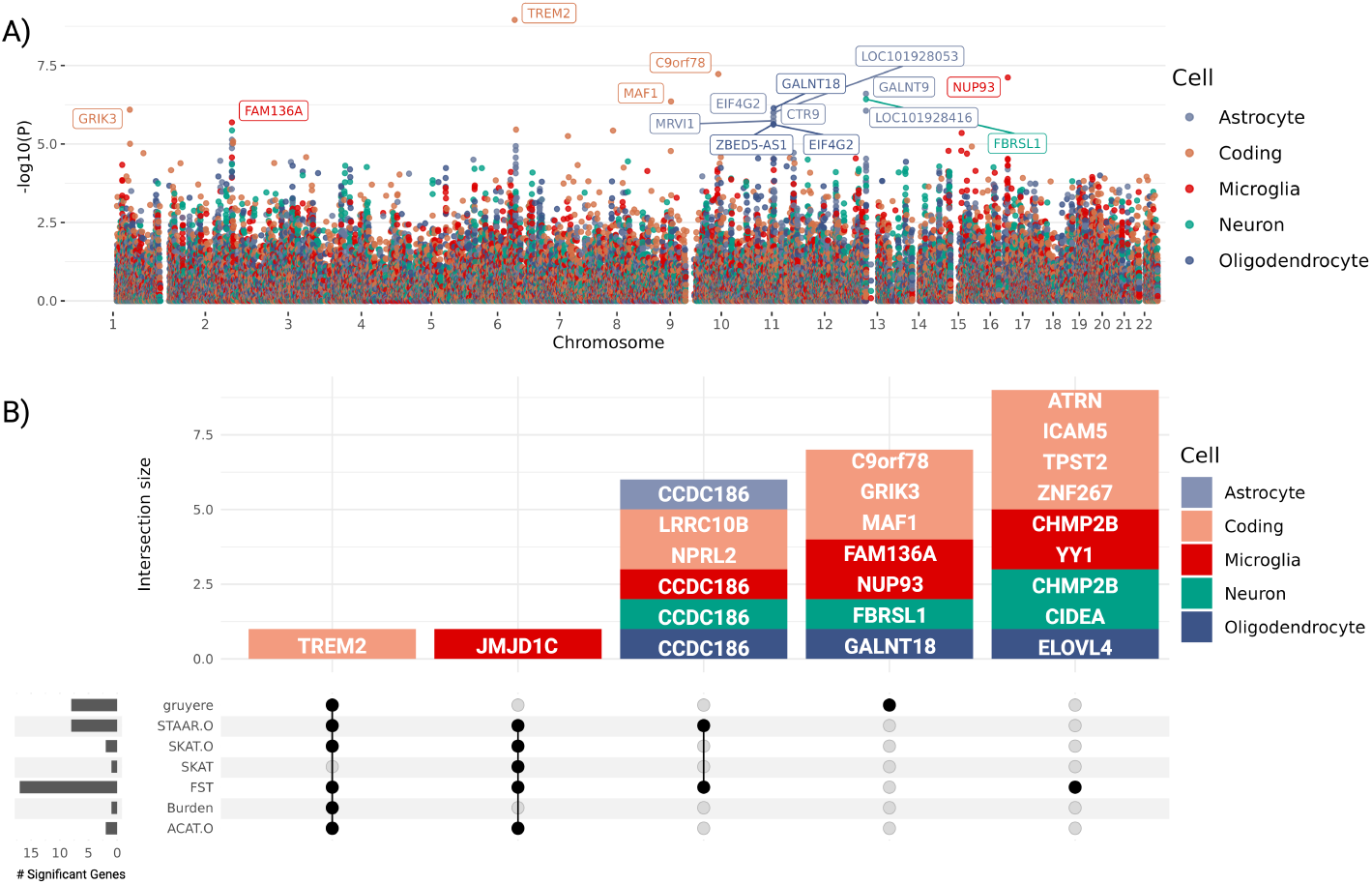
Top gruyere genes. **A)** Manhattan plot across cell types. The *Y* -axis shows *−* log_10_(*p*-value) for each gene and *X*-axis shows gene position. Each color is a cell type. `gruyere`-significant genes are labeled. **B)** Stacked upset plot of significant gene overlap across all tests after pruning for coregulation. Dark grey bars on left indicate total number of significant genes for each test. Vertically connected dots represent groups and corresponding bars indicate the number of overlapping significant genes identified for that group. Each bar is colored by cell type.

`gruyere` identified 12 non-coding RV associations across cell types with 2 in microglia, 6 in astrocytes, 1 in neurons, and 3 in oligodendrocytes. The most significant of these is *NUP93* (microglia), which, although not specifically linked to AD, is part of a group of nucleoporin (Nup) mutations associated with neurodegenerative disorders like AD [38]. Three significant genes, *GALNT9, FBRSL1*, and *LOC101928416*, are closely located on chromosome 12q24.33 and share over 80% of their ABC-predicted CREs, indicating that their associations are driven by the same set of RVs. Although variants can map to multiple genes in our model framework, making our analysis susceptible to coregulation, we are able to investigate and identify the specific CREs driving these associations. Of the overlapping promoters for these three genes, regions have higher ABC scores for *FBRSL1* (ABC = 0.20) compared to *GALNT9* (ABC = 0.04) and *LOC101928416* (ABC = 0.06), suggesting a stronger regulatory impact on *FBRSL1* and further isolating overlapping signal. *FBRSL1* (neuron), has not been linked to AD, but it presents a strong candidate gene for its distinctive neuronal expression profile and involvement in neurogenesis and transcriptional regulatory networks [39]. Multiple associations (*GALNT18, CTR9, EIF4G2, ZBED5-AS1, LOC101928053*, and *MRVI1*) specific to astrocyte and oligodendrocyte cell-types are coregulated in chromosome 11p15.4, and the strongest signal, *GALNT18*, has been connected to AD in more than one study [40, 41]. After pruning coregulated signals, `gruyere` identifies 8 significant genes.

We identify both known and novel AD-associated risk genes with `gruyere`. Significant `gruyere` genes are associated with increased gene expression across thirteen brain tissues found in GTEx (2-sample t-test *p* = 9.2 × 10^*−*31^, Supplemental Figure 3A) [42]. 5 of our 16 significant genes have expression QTLs in our microglia genomic atlas (isoMiGA) that colocalize with AD or Parkinson’s disease (PD) GWAS (Supplemental Figure 3B) [43–46]. Specifically, *TREM2* and *MAF1* have significant SNPs in a recent AD GWAS [44] while *FBRSL1, EIF4G2*, and *ZBED5-AS1* are significant in a large PD GWAS [45]. AD and PD have known genetic overlap, motivating QTL colocalization of both traits [47]. Finally, we compare `gruyere` *p-*values with the Alzheimer’s Disease Variant Portal (ADVP) catalog of 956 reported AD genes, finding that `gruyere` yields more significant *p-*values for ADVP versus non-ADVP genes (2-sample t-test *p* = 7.1 × 10^*−*9^, Supplemental Figure 3C) [48].

#### Comparing gruyere to existing methods

We compare pruned `gruyere` results with AD associations identified by a number of existing RV methods: burden test, sequence kernel association test (SKAT), optimal unified test (SKAT-O), functional score test (FST), aggregated Cauchy association test (ACAT), and variant-set test for association using annotation information (STAAR) [10, 25, 27, 49–51]. Burden, SKAT, SKAT-O, and ACAT-O tests do not include functional annotations, while FST and STAAR incorporate them (description in Supplemental Table 1 and detailed in Figure 1D for *TREM2* and microglia). We use the same set of functional annotations for FST and STAAR as for `gruyere`. We find that `gruyere` − *log*_10_(*p-*values) have the highest correlation with Burden tests (Pearson *R* = 0.86) and show moderate to high correlation with combination methods STAAR-O, SKAT-O, FST, and ACAT-O (Pearson *R* = 0.45 − 0.58) (Supplemental Figure 4A). The higher observed correlation with burden tests is expected, as `gruyere` also assumes unidirectional variant effects within a gene. We examine overlap of significant genes across all tests and find that there is minimal overlapping signal across methods (Figure 6B). Of the 16 (8 pruned) AD associations identified by `gruyere`, 15 (7 pruned) are unique to `gruyere`, while *TREM2* (coding) is detected across all tests but SKAT where it narrowly misses significance. In total, burden, SKAT, SKAT-O, and ACAT-O tests identify only two significant associations, highlighting the importance of including functional annotations, particularly for non-coding RV associations.

#### Integrating gruyere into omnibus tests

We combine `gruyere` *p-*values with existing methods using ACAT (Supplemental Figure 4B). Comparing *ACAT* (burden, SKAT, SKAT-O, FST, ACAT, STAAR) to *ACAT* (`gruyere`, burden, SKAT, SKAT-O, FST, ACAT, STAAR), we find that the inclusion of `gruyere` in omnibus tests boosts the number of significant associations from 12 to 16, adding *C9orf78, MAF1, NUP93* and *GALNT9*. There is no loss of power with this method, as all existing signals remain after including `gruyere`; we simply increase the total number of AD associations identified.

## 3 Methods

### 3.1 Data Overview

#### Whole-Genome Sequencing Data

We analyze the latest release of WGS data from the Alzheimer’s disease sequencing project (ADSP) consisting of 21,378 unrelated individuals over the age of 65 (7,966 cases, 13,412 controls) after QC [52**?**]. We follow a standard pipeline to QC WGS data. First, we combine phenotype information across multiple cohorts and remove genetically identical duplicates (IBD 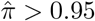) and technical replicate samples, selecting samples with the highest call rates. We prioritize phenotype information for individuals in family studies over case-control studies. Related individuals are removed using Kinship-based INference for Gwas (KING) [53], keeping AD cases where possible. In PLINK [54], we remove individuals with more than 10% genotype missingness, variants with less than 90% genotyping rate, and keep only biallelic variants with an observed *MAF* ≤ 0.05. Missing genotypes are imputed as the average observed MAF. For analysis, we randomly split samples into 80% train and 20% unseen test sets, stratifying by ancestry. ADSP samples are primarily of European (N = 9,133), African (N = 5,173) and Hispanic (5,059) ancestry, with smaller South Asian (N = 1,951) and East Asian (N = 62) groups.

#### Clinical Information

We use 30 available covariates in our model: sex, age, age^2^, age × sex, age × sex^2^, *APOE-e4* genotype, *APOE-e2* genotype, 10 ancestry principal components calculated from the 1000 Genomes Project [55], a common variant PRS [56], one-hot encoded sequencing platform (Illumina HiSeq 2000, HiSeqX, Nova Seq), one-hot encoded sequencing center (Illumina, USUHS, USUHS/Miami, NYGC, MEDGENOME, Baylor, Broad, WashU), and an intercept term (Figure 1A). Covariates are min-max scaled to a range of 0 to 1.

### 3.2 Proposed Bayesian rare variant model: gruyere

We develop `gruyere`, a hierarchy of per-gene GLMs (Supplemental Figure 1). We define our model jointly as

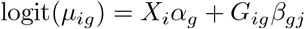

where *µ*_*ig*_ is the probability of AD for individual *i* associated with gene *g, X*_*i*_ is a vector of covariates, and *G*_*ig*_ is a vector of genotype dosages for each RV. Covariate coefficients *α*_*g*_ are modeled from prior,

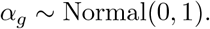

The key innovation in the `gruyere` model is the construction of per-variant genetic effects for gene *g*, or *β*_*gj*_ = (*β*_*gj*1_, …, *β*_*gjp*_)^*T*^, which is defined as the product of gene effects, transformed MAF, and weighted functional annotations. Of note, if a variant *j* is included in the RV set for both genes *g*_1_ and *g*_2_, the variant effect can differ for *β*_*g*1*j*_ and *β*_*g*2 *j*_. We define *β*_*gj*_ as

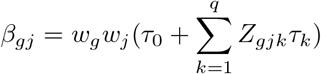

where *w*_*g*_ are gene importance weights, *w*_*j*_ are variant weights based on observed MAF as suggested by Wu et al.[49], *τ* are genome-wide annotation importance scores, and *Z*_*gjk*_ is a scaled functional annotation *k* for RV *j* and gene *g*. The variables are modeled as,

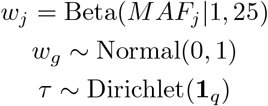

For each gene, we use a Bernoulli likelihood to sample *σ*(*X*_*i*_*α*_*g*_ + *G*_*ig*_*β*_*gj*_), and aggregate loss across each *g* ∈ *M*. Learned parameters are *α*_*g*_, *w*_*g*_, and *τ*. We select a Dirichlet prior for annotation weights *τ* to ensure identifiability between *τ* and *w*_*g*_. Without constraining *τ* to a fixed sum, *w*_*g*_ can be swapped for *w*_*g*_*/c* and *τ* for *cτ* for any positive constant *c* without changing the likelihood, leading to non-identifiability between gene and annotation weights.

We approximate the true posterior distribution for `gruyere` by minimizing the Kullback-Leibler (KL) divergence, which is equivalent to maximizing the Evidence Lower Bound (ELBO) [31]. To maximize the ELBO, we use SVI, implemented in the `pyro` probabilistic programming language [57]. We approximate the posterior distribution of latent variables *α*_*g*_ and *w*_*g*_ with mean field normal distributions (AutoNormal guide), while optimizing annotation weights *τ* as point estimates with a Delta distribution (AutoDelta guide). We apply the Adam optimizer, a learning rate of 0.1, train for 300 epochs, and draw 50 samples from the posterior to estimate standard deviations of the learned parameters. We explore different prior distributions for all key parameters.

Once global *τ* is learned, we streamline `gruyere` with a per-gene analysis. Holding *τ* fixed, our model simplifies to a logistic regression where only *α*_*g*_ and *w*_*g*_ are estimated. `gruyere` efficiently computes gene-level *p*-values using LRTs comparing a covariate-only regression to a combined covariate and genotype regression model:

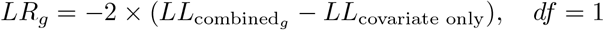

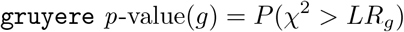

where *LL* are the log-likelihoods for each logistic regression. For each cell type, we use Bonferroni correction to define the *p-*value significance threshold:

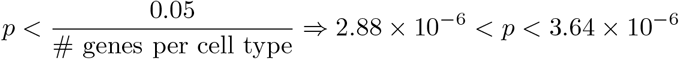

### 3.3 Cell-type specific RV gene set prediction using the ABC model

We calculate enhancer-gene connectivity using publicly available ATAC-seq and H3K27ac ChIP-seq data for human microglia, oligodendrocytes, astrocytes, and neurons [28]. We apply ABC to this data following the guidelines and default parameters provided at https://github.com/broadinstitute/ABC-Enhancer-Gene-Prediction. This involves first calling candidate peak regions for the ATAC-seq DNase hypersensitive sites (DHS) using MACS2 (peakExtendFromSummit = 250, nStrongestPeaks = 150000). Then we quantify enhancer activity as the geometric mean of the read counts of DHS and H3K27ac ChIP-seq in candidate enhancer regions. Lastly, we compute the ABC score using averaged Hi-C data (hic resolution = 5000) fit to the power-law model. The omics data is aligned to hg19, so we converted the ABC-predicted start and end positions of enhancers to hg38 for analysis. For each gene and separately each cell type, we aggregate all elements *E* for gene *G* that have an *ABC* ≥ 0.02 and extract RVs from within these regions to determine our cell-type specific non-coding RV gene sets.

### 3.4 Calculating functional annotation groups

We use a range of variant-level functional annotations primarily from the Whole Genome Sequencing Annotation database [58]. Annotations with greater than 5% missingness in our RVs are removed, resulting in 50 coding variant and 52 non-coding variant functional annotations listed in Supplemental Table 2. To reduce dimensionality of related annotations while accounting for their diverse biological effects, we apply non-negative matrix factorization (NMF) to summarize groups of related annotations, inspired by STAAR [10]. We use NMF to retain interpretable directional scaling of annotations. Based on correlation structure and a priori knowledge, we define six major functional categories – splicing, conservation, integrative deleterious predictions, brain-related Roadmap epigenetics, population-specific MAF, and enhancer activity [15, 59, 60]. Because the splicing annotation group is derived from four SpliceAI predictions that are not highly correlated and sparsely distributed, we instead use the maximum score for this category. We also include a binary LoF prediction calculated with Loss-Of-Function Transcript Effect Estimator (LOFTEE) [61] for coding variants along with an intercept term. All annotations are scaled between 0 and 1, where a larger value represents increased predicted variant function.

#### Deep Learning Delta Scores

For all four cell types, we include additional cell-type specific functional annotations: absolute maximum and absolute minimum TF delta scores derived from Enformer [14], a deep learning genomics model. We calculate variant delta scores for 5,318 functional genomics assays. The Enformer model predicts read counts (in 128 BP bins) of these assays as a function of 196,608 BP input DNA sequence. For a particular variant, we compare the model output of the reference sequence, centered around the variant position, with the output of the alternative sequence which replaces the reference allele with the alternative allele. For a particular genomics assay, the delta score is the difference between the sum of reference sequence predictions for the middle 32 bins and the sum of the alternate sequence predictions for the same bins. We normalize these scores by first calculating the delta scores for the approximately 18 million variants from the UK Biobank cohort used in PolyFun [62, 63], and then Z-score normalize each assay according based on this collection of variants. We apply this normalization to the delta scores used in our analysis. We aggregate delta scores to determine composite maximum and minimum predictions for each variant, highlighting the delta scores of only the enriched TFs within each of the four cell types ([28]). For microglia non-coding variant sets, we additionally use delta scores for 4 epigenetic marks (H3K4me3, H3K27ac, H3K27me3, and DNASE) for monocytes, a proxy for microglia.

### 3.5 Data Simulation

We use simulations to evaluate `gruyere` performance. We randomly sample values for each parameter and use these simulated variables in the GLM framework, logit(*µ*_*ig*_) = *X*_*i*_*α*_*g*_ + *G*_*ig*_*β*_*g*_. The real ADSP genotypes *G*_*ig*_, covariates *X*_*i*_, and functional annotations *Z*_*gjk*_ along with simulated parameters 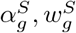, and *τ* ^*S*^, generate simulated phenotypes 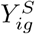. Simulations are restricted to a maximum estimated heritability of 30% to realistically evaluate complex diseases. For each simulation, we draw `gruyere` parameters from the following distributions:

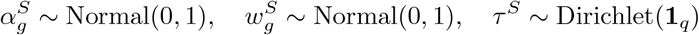

We define *β*_*gj*_ in the same way, simply using simulated variables:

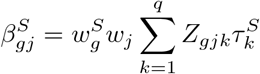

Using this combination of true data and simulated parameters, we sample synthetic phenotypes 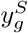 from a Bernoulli distribution. We perform 100 simulations on *M* = 500 randomly selected genes. In general, we sample `gruyere` parameters from the same distribution that they are learned. We have also tested model performance when simulated data comes from a different distribution than its learned counterpart (e.g. sampling *w*_*g*_ from a Normal distribution in simulations but fitting from a Gamma prior).

## 4 Discussion

We develop `gruyere`, a functionally-informed RV association test that fits a hierarchy of Bayesian GLMs to estimate genome-wide functional annotation importance, gene effects, and covariate coefficients. `gruyere` builds upon existing RV methods with two key advancements: 1) a genome-wide approach that enables trait-specific weighting of functional annotations, and 2) a flexible, powerful and calibrated probabilistic framework that estimates uncertainty. We incorporate an innovative methodology for analyzing RVs in the non-coding genome. Using the Activity-by-Contact model, we predict cell-type-specific enhancer-gene connectivity from chromatin state and conformation data, aggregating predictions by gene to define interpretable non-coding RV testing regions. We use in silico mutagenesis under state-of-the-art deep learning models of pre (SpliceAI) and post (Enformer) transcription gene regulation to predict RV effects. Simulation analyses validate `gruyere` and show it is able to recover ground truth parameters across diverse model specifications and even for realistically low heritability.

We apply `gruyere`, along with a number of established RV association tools, to the most recent WGS release from ADSP. Our analysis identifies both known (e.g., *TREM2*) and novel (e.g., *NUP93*) candidate AD genes. Specifically, `gruyere` uniquely identifies 15 genes, of which *C9orf78, MAF1, NUP93* and *GALNT9* remain significant in aggregated Cauchy tests. Our analysis additionally provides an improved understanding of AD-relevant functional annotations. `gruyere` confirms the expectation that LoF is the most informative annotation for coding variants, but additionally finds deep learning-based predictions for splicing, TF binding and chromatin state are highly predictive of functional non-coding RVs.

We use ancestry principal components as covariates to account for population diversity, but one area for future work would be integrating a random effect term to better account for relatedness and population structure [64]. Another possible extension to `gruyere` would be incorporating gene-level features as priors [65]. While we focus our analysis on AD, `gruyere` can be applied to any complex disease with sufficient WGS data. As the quality of functional annotations continues to improve, `gruyere` will become an increasingly valuable tool for identifying disease-associated genes and annotations.

## Supplementary information

Supplementary Figures 1-4; Supplementary Table 1-2; Supplementary Methods; Supplementary Acknowledgements; Supplementary Data: Gene *p-*values for each cell-type.

## Acknowledgements

Research reported in this paper was supported by Alzheimer’s Disease Sequencing Project of the National Institutes of Health under award number 5 U01 AG068880-02. The content is solely the responsibility of the authors and does not necessarily represent the official views of the National Institutes of Health. We are thankful to Tulsi Patel, Edoardo Marcora, Alison Goate, and Iuliana Ionita-Laza for helpful feedback and discussions.

## Declarations

### Funding

Research reported in this paper was supported by Alzheimer’s Disease Sequencing Project of the National Institutes of Health under award number 5 U01 AG068880-02.

### Declaration of interests

T.R. served as a scientific advisor for Merck and serves as a consultant for Curie.Bio.

### Ethics approval and consent to participate

Not applicable

### Consent for publication

Not applicable

### Data availability

This paper uses the ADSP Release 4 WGS data and AD phenotype data.

### Materials availability

Not applicable

### Code availability

Code and details for running `gruyere` is available on GitHub: https://github.com/daklab/gruyere

### Author contribution

D.A.K. conceived and supervised the project. A.D. and D.A.K. developed the methods. A.D. and D.A.K. wrote the manuscript. A.D. wrote the software code and performed the analyses. All authors read, reviewed, and approved the final manuscript.

### Editorial Policies for

Springer journals and proceedings: https://www.springer.com/gp/editorial-policies

Nature Portfolio journals: https://www.nature.com/nature-research/editorial-policies

*Scientific Reports*: https://www.nature.com/srep/journal-policies/editorial-policies

BMC journals: https://www.biomedcentral.com/getpublished/editorial-policies

## Notes

### Competing Interest Statement

The authors have declared no competing interest.

### Funding Statement

This study was funded by Alzheimer's Disease Sequencing Project of the National Institutes of Health under award number 5 U01 AG068880-02.

### Author Declarations

The IRB of Columbia University gave ethical approval for this work.

### Summary of Updates

Updated declaration of interest. We missed this on the initial submission.

## References

[1] Sims, R., Hill, M., Williams, J.: The multiplex model of the genetics of alzheimer’s disease. Nat. Neurosci. 23, 311–322 (2020) 10.1038/s41593-020-0599-5

[2] Andrews, S.J., Renton, A.E., Fulton-Howard, B., Podlesny-Drabiniok, A., Marcora, E., Goate, A.M.: The complex genetic architecture of alzheimer’s disease: novel insights and future directions 90, 104511 (2023) 10.1016/j.ebiom.2023.104511

[3] Auer, P.L., Lettre, G.: Rare variant association studies: considerations, challenges and opportunities 7, 16 (2015) 10.1186/s13073-015-0138-2

[4] Wainschtein, P., Jain, D., Zheng, Z., TOPMed Anthropometry Working Group, NHLBI Trans-Omics for Precision Medicine (TOPMed) Consortium, Cupples, L.A., Shadyab, A.H., McKnight, B., Shoemaker, B.M., Mitchell, B.D., Psaty, B.M., Kooperberg, C., et al.: Assessing the contribution of rare variants to complex trait heritability from whole-genome sequence data 54, 263–273 (2022) 10.1038/s41588-021-00997-7

[5] Kosmicki, J.A., Churchhouse, C.L., Rivas, M.A., Neale, B.M.: Discovery of rare variants for complex phenotypes. Hum. Genet. 135, 625–634 (2016) 10.1007/s00439-016-1679-1

[6] Fiziev, P.P., McRae, J., Ulirsch, J.C., Dron, J.S., Hamp, T., Yang, Y., Wainschtein, P., Ni, Z., Schraiber, J.G., Gao, H., Cable, D., Field, Y., Aguet, F., Fasnacht, M., Metwally, A., Rogers, J., Marques-Bonet, T., Rehm, H.L., O’Donnell-Luria, A., Khera, A.V., Farh, K.K.-H.: Rare penetrant mutations confer severe risk of common diseases. Science 380, 1131 (2023) 10.1126/science.abo1131

[7] Hoogmartens, J., Cacace, R., Van Broeckhoven, C.: Insight into the genetic etiology of alzheimer’s disease: A comprehensive review of the role of rare variants 13, 12155 (2021) 10.1002/dad2.12155

[8] Li, Z., Li, X., Zhou, H., Gaynor, S.M., Selvaraj, M.S., Arapoglou, T., Quick, C., Liu, Y., Chen, H., Sun, R., et al.: A framework for detecting noncoding rare-variant associations of large-scale whole-genome sequencing studies 19, 1599–1611 (2022) 10.1038/s41592-022-01640-x

[9] Lee, S., Abecasis, G.R., Boehnke, M., Lin, X.: Rare-variant association analysis: study designs and statistical tests. Am. J. Hum. Genet. 95, 5–23 (2014) 10.1016/j.ajhg.2014.06.009

[10] Li, X., Li, Z., Zhou, H., Gaynor, S.M., Liu, Y., Chen, H., Sun, R., Dey, R., Arnett, D.K., Aslibekyan, S., et al.: Dynamic incorporation of multiple in silico functional annotations empowers rare variant association analysis of large whole-genome sequencing studies at scale 52, 969–983 (2020) 10.1038/s41588-020-0676-4

[11] Clarke, B., Holtkamp, E., Öztürk, H., Mück, M., Wahlberg, M., Meyer, K., Munzlinger, F., Brechtmann, F., Hölzlwimmer, F.R., Gagneur, J., Stegle, O.: Integration of variant annotations using deep set networks boosts rare variant association genetics, 2023–0712548506 (2023) 10.1101/2023.07.12.548506

[12] Wagner, N., Çelik, M.H., Hölzlwimmer, F.R., Mertes, C., Prokisch, H., Yépez, V.A., Gagneur, J.: Aberrant splicing prediction across human tissues 55, 861–870 (2023) 10.1038/s41588-023-01373-3

[13] Zeng, T., Li, Y.I.: Predicting RNA splicing from DNA sequence using pangolin 23, 103 (2022) 10.1186/s13059-022-02664-4

[14] Avsec,, Agarwal, V., Visentin, D., Ledsam, J.R., Grabska-Barwinska, A., Taylor, K.R., Assael, Y., Jumper, J., Kohli, P., Kelley, D.R.: Effective gene expression prediction from sequence by integrating long-range interactions. Nat. Methods 18, 1196–1203 (2021) 10.1038/s41592-021-01252-x

[15] Pollard, K.S., Hubisz, M.J., Rosenbloom, K.R., Siepel, A.: Detection of non-neutral substitution rates on mammalian phylogenies 20, 110–121 (2010) 10.1101/gr.097857.109

[16] Ionita-Laza, I., McCallum, K., Xu, B., Buxbaum, J.D.: A spectral approach integrating functional genomic annotations for coding and noncoding variants 48, 214–220 (2016) 10.1038/ng.3477

[17] Skene, N.G., Grant, S.G.N.: Identification of vulnerable cell types in major brain disorders using single cell transcriptomes and expression weighted cell type enrichment 10, 16 (2016) 10.3389/fnins.2016.00016

[18] Li, Z., Li, X., Liu, Y., Shen, J., Chen, H., Zhou, H., Morrison, A.C., Boerwinkle, E., Lin, X.: Dynamic scan procedure for detecting rare-variant association regions in whole-genome sequencing studies. Am. J. Hum. Genet. 104, 802–814 (2019) 10.1016/j.ajhg.2019.03.002

[19] Sey, N.Y.A., Hu, B., Mah, W., Fauni, H., McAfee, J.C., Rajarajan, P., Brennand, K.J., Akbarian, S., Won, H.: A computational tool (H-MAGMA) for improved prediction of brain-disorder risk genes by incorporating brain chromatin interaction profiles. Nat. Neurosci. 23, 583–593 (2020) 10.1038/s41593-020-0603-0

[20] Ma, S., Dalgleish, J., Lee, J., Wang, C., Liu, L., Gill, R., Buxbaum, J.D., Chung, W.K., Aschard, H., Silverman, E.K., Cho, M.H., He, Z., Ionita-Laza, I.: Powerful gene-based testing by integrating long-range chromatin interactions and knockoff genotypes 118 (2021) 10.1073/pnas.2105191118

[21] Zhang, S., Moll, T., Rubin-Sigler, J., Tu, S., Li, S., Yuan, E., Liu, M., Butt, A., Harvey, C., et al.: Deep learning modeling of rare noncoding genetic variants in human motor neurons definesCCDC146 as a therapeutic target for ALS (2024) 10.1101/2024.03.30.24305115

[22] Fulco, C.P., Nasser, J., Jones, T.R., Munson, G., Bergman, D.T., Subramanian, V., Grossman, S.R., Anyoha, R., Doughty, B.R., Patwardhan, et al.: Activity-by-contact model of enhancer-promoter regulation from thousands of CRISPR perturbations. Nat. Genet. 51, 1664–1669 (2019) 10.1038/s41588-019-0538-0

[23] Hoffman, M.D., Blei, D.M., Wang, C., Paisley, J.: Stochastic variational inference. J. Mach. Learn. Res. (2013)

[24] Jaganathan, K., Kyriazopoulou Panagiotopoulou, S., McRae, J.F., Darbandi, S.F., Knowles, D., Li, Y.I., Kosmicki, J.A., Arbelaez, J., Cui, W., Schwartz, G.B., Chow, E.D., Kanterakis, E., Gao, H., Kia, A., Batzoglou, S., Sanders, S.J., Farh, K.K.-H.: Predicting splicing from primary sequence with deep learning 176, 535–54824 (2019) 10.1016/j.cell.2018.12.015

[25] Madsen, B.E., Browning, S.R.: A groupwise association test for rare mutations using a weighted sum statistic. PLoS Genet. 5, 1000384 (2009) 10.1371/journal.pgen.1000384

[26] Morgenthaler, S., Thilly, W.G.: A strategy to discover genes that carry multiallelic or mono-allelic risk for common diseases: a cohort allelic sums test (CAST) 615, 28–56 (2007) 10.1016/j.mrfmmm.2006.09.003

[27] He, Z., Xu, B., Lee, S., Ionita-Laza, I.: Unified sequence-based association tests allowing for multiple functional annotations and meta-analysis of noncoding variation in metabochip data 101, 340–352 (2017) 10.1016/j.ajhg.2017.07.011

[28] Nott, A., Holtman, I.R., Coufal, N.G., Schlachetzki, J.C.M., Yu, M., Hu, R., Han, C.Z., Pena, M., Xiao, J., Wu, et al.: Brain cell type-specific enhancer-promoter interactome maps and disease-risk association. Science 366, 1134–1139 (2019) 10.1126/science.aay0793

[29] Dempster, E.R., Lerner, I.M.: Heritability of threshold characters 35, 212–236 (1950) 10.1093/genetics/35.2.212

[30] Zhou, X., Chen, Y., Ip, F.C.F., Jiang, Y., Cao, H., Lv, G., Zhong, H., Chen, J., Ye, T., Chen, et al.: Deep learning-based polygenic risk analysis for alzheimer’s disease prediction. Commun. Med. (Lond.) 3, 49 (2023) 10.1038/s43856-023-00269-x

[31] Blei, D.M., Kucukelbir, A., McAuliffe, J.D.: Variational inference: A review for statisticians. J. Am. Stat. Assoc. 112, 859–877 (2017) 10.1080/01621459.2017.1285773

[32] Gopinath, A., Collins, A., Khoshbouei, H., Streit, W.J.: Microglia and other myeloid cells in central nervous system health and disease. J. Pharmacol. Exp. Ther. 375, 154–160 (2020) 10.1124/jpet.120.265058

[33] Gratuze, M., Leyns, C.E.G., Holtzman, D.M.: New insights into the role of TREM2 in alzheimer’s disease. Mol. Neurodegener. 13, 66 (2018) 10.1186/s13024-018-0298-9

[34] Han, Y., Chen, K., Yu, H., Cui, C., Li, H., Hu, Y., Zhang, B., Li, G.: Maf1 loss regulates spinogenesis and attenuates cognitive impairment in alzheimer’s disease. Brain 147, 2128–2143 (2024) 10.1093/brain/awae015

[35] Bottero, V., Potashkin, J.A.: Meta-analysis of gene expression changes in the blood of patients with mild cognitive impairment and alzheimer’s disease dementia. Int. J. Mol. Sci. 20, 5403 (2019) 10.3390/ijms20215403

[36] Izadi, F., Soheilifar, M.H.: Exploring potential biomarkers underlying pathogenesis of alzheimer’s disease by differential co-expression analysis 10, 233–241 (2018)

[37] Xiao, B., Kuang, Z., Zhang, W., Hang, J., Chen, L., Lei, T., He, Y., Deng, C., Li, W., Lu, J., Qu, J., et al.: Glutamate ionotropic receptor kainate type subunit 3 (GRIK3) promotes epithelial-mesenchymal transition in breast cancer cells by regulating SPDEF/CDH1 signaling 58, 1314–1323 (2019) 10.1002/mc.23014

[38] Spead, O., Zaepfel, B.L., Rothstein, J.D.: Nuclear pore dysfunction in neurodegeneration. Neurotherapeutics 19, 1050–1060 (2022) 10.1007/s13311-022-01293-w

[39] Bukvic, N., De Rinaldis, M., Chetta, M., Trabacca, A., Bassi, M.T., Marsano, R.M., Holoubkova, L., Rivieccio, M., Oro, M., Resta, et al.: De novo pathogenic variant in FBRSL1, non OMIM gene paralogue AUTS2, causes a novel recognizable syndromic manifestation with intellectual disability; an additional patient and review of the literature. Genes (Basel) 15, 826 (2024) 10.3390/genes15070826

[40] Mishra, R., Li, B.: The application of artificial intelligence in the genetic study of alzheimer’s disease. Aging Dis. 11, 1567–1584 (2020) 10.14336/AD.2020.0312

[41] Lutz, M.W., Sprague, D., Barrera, J., Chiba-Falek, O.: Shared genetic etiology underlying alzheimer’s disease and major depressive disorder. Transl. Psychiatry 10, 88 (2020) 10.1038/s41398-020-0769-y

[42] GTEx Consortium: The GTEx consortium atlas of genetic regulatory effects across human tissues 369, 1318–1330 (2020) 10.1126/science.aaz1776

[43] Humphrey, J., Brophy, E., Kosoy, R., Zeng, B., Coccia, E., Mattei, D., Ravi, A., Efthymiou, A.G., Navarro, E., Muller, B.Z., et al.: Long-read RNA-seq atlas of novel microglia isoforms elucidates disease-associated genetic regulation of splicing. medRxiv, 2023–120123299073 (2023) 10.1101/2023.12.01.23299073

[44] Bellenguez, C., Küçükali, F., Jansen, I.E., Kleineidam, L., Moreno-Grau, S., Amin, N., Naj, A.C., Campos-Martin, R., Grenier-Boley, B., Andrade, et al.: New insights into the genetic etiology of alzheimer’s disease and related dementias. Nat. Genet. 54, 412–436 (2022) 10.1038/s41588-022-01024-z

[45] Lambert, J.C., Ibrahim-Verbaas, C.A., Harold, D., Naj, A.C., Sims, R., Bellenguez, C., DeStafano, A.L., Bis, J.C., Beecham, G.W., Grenier-Boley, B., Russo, et al.: Meta-analysis of 74,046 individuals identifies 11 new susceptibility loci for alzheimer’s disease. Nat. Genet. 45, 1452–1458 (2013) 10.1038/ng.2802

[46] Nalls, M.A., Blauwendraat, C., Vallerga, C.L., Heilbron, K., Bandres-Ciga, S., Chang, D., Tan, M., Kia, D.A., Noyce, A.J., et al.: Identification of novel risk loci, causal insights, and heritable risk for parkinson’s disease: a meta-analysis of genome-wide association studies. Lancet Neurol. 18, 1091–1102 (2019) 10.1016/S1474-4422(19)30320-5

[47] Desikan, R.S., Schork, A.J., Wang, Y., Witoelar, A., Sharma, M., McEvoy, L.K., Holland, D., Brewer, J.B., Chen, C.-H., Thompson, W.K., et al.: Genetic overlap between alzheimer’s disease and parkinson’s disease at the MAPT locus. Mol. Psychiatry 20, 1588–1595 (2015) 10.1038/mp.2015.6

[48] Kuksa, P.P., Liu, C.-L., Fu, W., Qu, L., Zhao, Y., Katanic, Z., Clark, K., Kuzma, A.B., Ho, P.-C., et al.: Alzheimer’s disease variant portal: A catalog of genetic findings for alzheimer’s disease. J. Alzheimers. Dis. 86, 461–477 (2022) 10.3233/JAD-215055

[49] Wu, M.C., Lee, S., Cai, T., Li, Y., Boehnke, M., Lin, X.: Rare-variant association testing for sequencing data with the sequence kernel association test. Am. J. Hum. Genet. 89, 82–93 (2011) 10.1016/j.ajhg.2011.05.029

[50] Lee, S., Emond, M.J., Bamshad, M.J., Barnes, K.C., Rieder, M.J., Nickerson, D.A., NHLBI GO Exome Sequencing Project—ESP Lung Project Team, Christiani, D.C., Wurfel, M.M., Lin, X.: Optimal unified approach for rare-variant association testing with application to small-sample case-control whole-exome sequencing studies 91, 224–237 (2012) 10.1016/j.ajhg.2012.06.007

[51] Liu, Y., Chen, S., Li, Z., Morrison, A.C., Boerwinkle, E., Lin, X.: ACAT: A fast and powerful p value combination method for rare-variant analysis in sequencing studies 104, 410–421 (2019) 10.1016/j.ajhg.2019.01.002

[52] Beecham, G.W., Bis, J.C., Martin, E.R., Choi, S.-H., DeStefano, A.L., Duijn, C.M., Fornage, M., Gabriel, S.B., Koboldt, D.C., Larson, D.E., et al.: The alzheimer’s disease sequencing project: Study design and sample selection. Neurol Genet 3, 194 (2017) 10.1212/NXG.0000000000000194

[53] Manichaikul, A., Mychaleckyj, J.C., Rich, S.S., Daly, K., Sale, M., Chen, W.-M.: Robust relationship inference in genome-wide association studies 26, 2867–2873 10.1093/bioinformatics/btq559

[54] Purcell, S., Neale, B., Todd-Brown, K., Thomas, L., Ferreira, M.A.R., Bender, D., Maller, J., Sklar, P., Bakker, P.I.W., Daly, M.J., Sham, P.C.: PLINK: a tool set for whole-genome association and population-based linkage analyses 81, 559–575 (2007) 10.1086/519795

[55] Siva, N.: 1000 genomes project 26, 256 (2008) 10.1038/nbt0308-256b

[56] Ge, T., Chen, C.-Y., Ni, Y., Feng, Y.-C.A., Smoller, J.W.: Polygenic prediction via bayesian regression and continuous shrinkage priors. Nat. Commun. 10, 1776 (2019) 10.1038/s41467-019-09718-5

[57] Eli, B., Jonathan, P.C., Martin, J., Fritz, O., Neeraj, P., Theofanis, K., Rohit, S., Paul, S., Paul, H., Noah, D.G.: Pyro: Deep universal probabilistic programming. arXiv [cs.LG] (2018) 10.48550/arXiv.1810.09538

[58] Liu, X., White, S., Peng, B., Johnson, A.D., Brody, J.A., Li, A.H., Huang, Z., Carroll, A., Wei, P., Gibbs, R., Klein, R.J., Boerwinkle, E.: WGSA: an annotation pipeline for human genome sequencing studies 53, 111–112 (2016) 10.1136/jmedgenet-2015-103423

[59] Bernstein, B.E., Stamatoyannopoulos, J.A., Costello, J.F., Ren, B., Milosavljevic, A., Meissner, A., Kellis, M., Marra, M.A., Beaudet, A.L., Ecker, J.R., et al.: The NIH roadmap epigenomics mapping consortium 28, 1045–1048 (2010) 10.1038/nbt1010-1045

[60] Chen, S., Francioli, L.C., Goodrich, J.K., Collins, R.L., Kanai, M., Wang, Q., Alföldi, J., Watts, N.A., Vittal, C., Gauthier, L.D., et al.: A genomic mutational constraint map using variation in 76,156 human genomes. Nature 625, 92–100 (2024) 10.1038/s41586-023-06045-0

[61] Karczewski, K.J., Francioli, L.C., Tiao, G., Cummings, B.B., Alföldi, J., Wang, Q., Collins, R.L., Laricchia, K.M., Ganna, A., Birnbaum, D.P., Gauthier, et al.: The mutational constraint spectrum quantified from variation in 141,456 humans. Nature 581, 434–443 (2020) 10.1038/s41586-020-2308-7

[62] Weissbrod, O., Hormozdiari, F., Benner, C., Cui, R., Ulirsch, J., Gazal, S., Schoech, A.P., Geijn, B., Reshef, Y., Márquez-Luna, C., O’Connor, L., Pirinen, M., Finucane, H.K., Price, A.L.: Functionally informed fine-mapping and polygenic localization of complex trait heritability. Nat Genet 52, 1355–1363 (2020) 10.1038/s41588-020-00735-5

[63] Szustakowski, J.D., Balasubramanian, S., Kvikstad, E., Khalid, S., Bronson, P.G., Sasson, A., Wong, E., Liu, D., Wade Davis, J., Haefliger, et al.: Advancing human genetics research and drug discovery through exome sequencing of the UK biobank. Nat Genet 53, 942–948 (2021) 10.1038/s41588-021-00885-0

[64] Zhou, W., Zhao, Z., Nielsen, J.B., Fritsche, L.G., LeFaive, J., Gagliano Taliun, S.A., Bi, W., Gabrielsen, M.E., Daly, M.J., Neale, B.M., et al.: Scalable generalized linear mixed model for region-based association tests in large biobanks and cohorts 52, 634–639 (2020) 10.1038/s41588-020-0621-6

[65] Londhe, S., Lindner, J., Chen, Z., Holtkamp, E., Hölzlwimmer, F.R., Casale, F.P., Brechtmann, F., Gagneur, J.: Functional gene embeddings improve rare variant polygenic risk scores. bioRxiv, 2024–0722604535 (2024) 10.1101/2024.07.22.604535

